# Developing Del2Phen: a novel phenotype description tool for chromosome deletions

**DOI:** 10.1101/2024.10.21.24315854

**Authors:** Eleana Rraku, Tyler D. Medina, Conny M.A. van Ravenswaaij-Arts, Mariska K. Slofstra, Morris A. Swertz, Trijnie Dijkhuizen, Lennart F. Johansson, Aafke Engwerda

**Affiliations:** University of Groningen, University Medical Center Groningen, Department of Genetics, Groningen, the Netherlands; Department of Human Genetics, Amsterdam University Medical Center, University of Amsterdam, the Netherlands; University of Galway, School of Mathematical and Statistical Sciences, Galway, Ireland; University of Groningen, University Medical Center Groningen, Groningen, Genomics Coordination Center, the Netherlands; Autism Team Northern-Netherlands, Jonx/Lentis Psychiatric Institute, Groningen, the Netherlands

**Keywords:** chromosome 6, chromosome disorder, tool, phenotype information, Chromosome 6 Project, parents

## Abstract

Information on the health-related consequences of rare chromosome disorders is often limited, posing challenges for both patients and their families. The Chromosome 6 Project aims to bridge this knowledge gap for structural aberrations involving chromosome 6 by providing parents of affected children with information on the expected phenotypes of their child. To achieve this, detailed phenotype and genotype data is collected directly from parents worldwide and supplemented with data from literature reports, resulting thus far in a dataset of over 500 individuals. This comprehensive data pool was used to develop Del2Phen, a software tool introduced in this paper that generates aberration-specific phenotype information for chromosome disorders. Del2Phen identifies individuals with a deletion or duplication similar to that of a new patient (index) and produces a clinical description for the index based on phenotypic features observed in these genotypically similar individuals. Genotypic similarity is determined using existing knowledge on the haploinsufficiency effect of genes and established gene-phenotype relationships. The optimal genotypic similarity parameters for chromosome 6 deletions were evaluated, which lead to thorough and reliable clinical descriptions based on sufficiently large groups of individuals with highly similar deletions. Although currently applied to and optimized for chromosome 6 deletions, Del2Phen can be used on deletions involving other chromosomes and is easily adapted for use on duplications, provided sufficient data is available. Del2Phen can already be used to expedite data analysis for chromosome disorders, thus aiding healthcare professionals in providing clinical care. Lastly, this tool will also be integrated into an interactive website aimed at parents of children with a chromosome 6 aberration, providing them with essential health information in an efficient and timely manner.

## 1. Introduction

Information on the phenotypic consequences of rare chromosome aberrations, including deletions and duplications, is often scarce. This leads to parental uncertainty about their child’s future, and knowledge about potential health issues has been shown to be crucial for parental coping.^1,2^ Better availability of such information would also help healthcare providers deliver appropriate care to affected individuals.

Studying the clinical manifestations of structural chromosome aberrations is challenging for several reasons. The rareness of these aberrations and the geographic spread of affected individuals means researchers must primarily rely on data from literature reports and online databases, both of which often provide incomplete information. Additionally, most structural aberrations (unless inherited) are unique in location, size and genetic content, making it difficult to identify genotypically similar individuals to compare and link to a phenotypic outcome. Lastly, determining the clinical effect of a chromosome abnormality is complex even for highly overlapping aberrations since the resulting phenotype is not based simply on the sum of all affected genes.

To address this knowledge gap, the Chromosome 6 Project (www.chromosome6.org) was established with a focus on structural aberrations affecting chromosome 6. This parent-driven research project aims to provide parents of affected individuals with detailed information on the possible clinical consequences of their child’s aberration. The Chromosome 6 Project uses social media to reach and recruit new participants and collects detailed phenotype and genotype information directly from parents of affected children.^3^ This information, supplemented with data extracted from literature reports and stored in the secure Chromosome 6 database, has been used to clinically characterise deletions in various chromosome 6 regions.^3–6^ Furthermore, a study conducted within the scope of this project has demonstrated that parent-derived phenotype information is suitable for clinical descriptions of rare chromosome disorders.^7^

Building upon a successful participant recruitment and data collection strategy, we can now proceed with the next steps in the project: automating and optimising the way genotype and phenotype data is analysed, so that clinical information is readily available to parents and clinicians. In the present study, we introduce Del2Phen, a software tool that produces tailored phenotype descriptions for chromosome aberrations using the clinical features of genotypically similar individuals. Here, we also evaluate the optimal genotypic similarity parameters for chromosome 6 deletions. Since these are rare and unique aberrations, we ascertained the best way for Del2Phen to group a sufficient number of genotypically similar individuals to produce a reliable phenotype description. Even though Del2Phen was developed using data on chromosome 6 deletions, it can be applied to deletions involving other chromosomes and is easily adaptable for use on duplications.

## 2. Materials and Methods

Del2Phen is a command-line Python 3 tool (installable from Github (https://github.com/Chromosome-6-Project/Del2Phen) or PyPI (pypi.org/project/del2phen) that uses existing genotype and phenotype data to generate a phenotype description tailored to the deletion of a new patient (index) (Figure 1). The tool identifies individuals with a deletion similar to that of the index and produces a phenotype description based on the clinical features of these individuals.

**Fig. 1:**
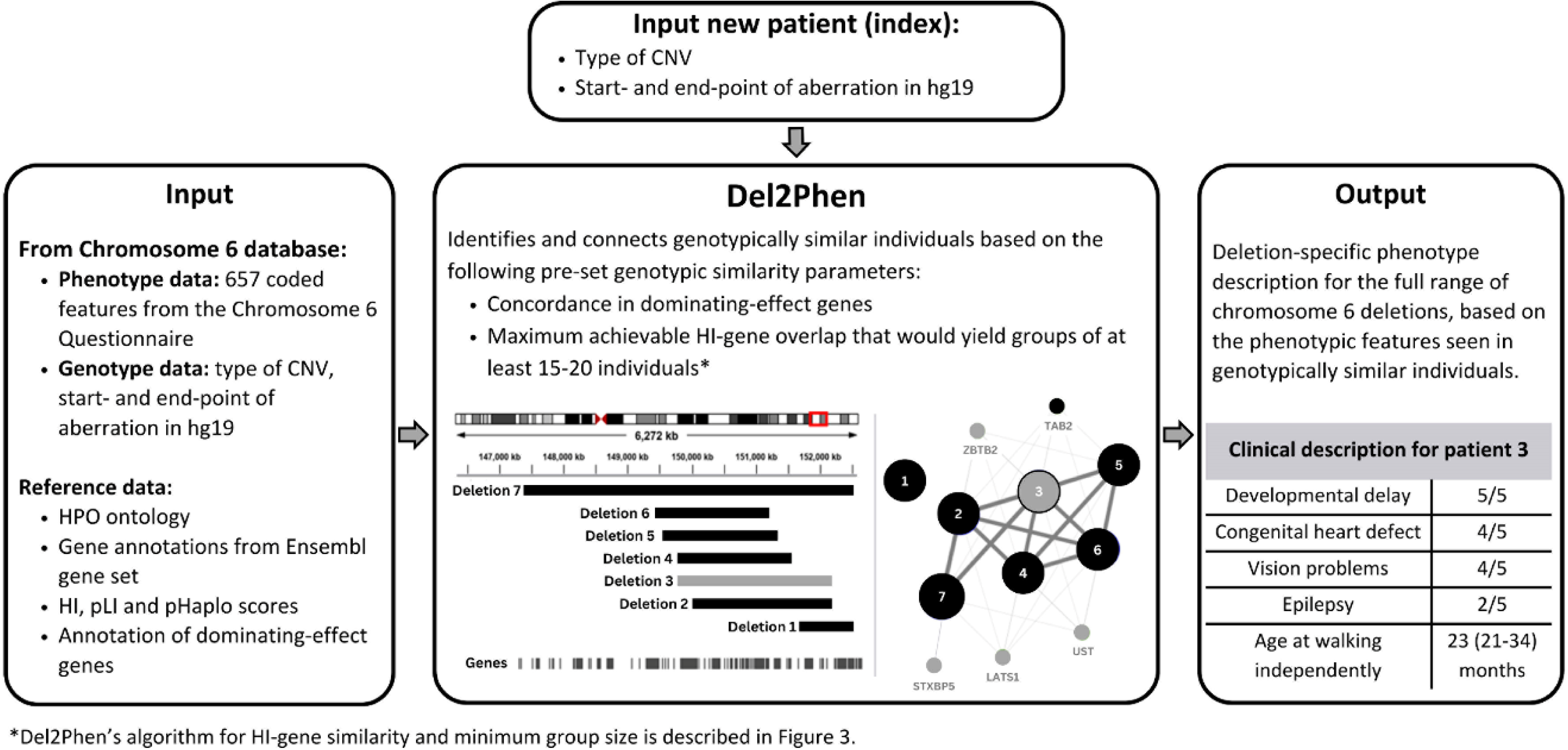
Schematic representation of Del2Phen. The tool uses existing genotype and phenotype data as input, in this study this is data stored in the Chromosome 6 database (left panel). Based on pre-set genotypic similarity parameters, Del2Phen identifies individuals with a similar deletion to that of an index patient (indicated in grey in middle panel). Deletions are identified as similar based on concordance in dominating-effect gene content and the maximum achievable HI-gene similarity that yields groups that exceed a certain size (following the algorithm developed in this study and depicted in Figure 3). Details on the definitions of HI- and dominating-effect genes are provided in Materials and Methods 2.1. Patients identified by the tool as genotypically similar to the index are grouped together, and their phenotypes are used to give a phenotype description for the index patient (right panel).

Genotypic similarity is determined based on pre-set parameters (see Materials and Methods 2.2). In this study, we applied and optimized Del2Phen for use on chromosome 6 deletions using data in the Chromosome 6 database. We determined the optimal genotypic similarity parameters for chromosome 6 deletions, which produce thorough and accurate clinical descriptions based on sufficiently large groups that share a high degree of genotypic similarity.

### 2.1. Del2Phen input and annotations

For this study, Del2Phen uses genotype and phenotype data stored in the Chromosome 6 database as input (Figure 1). This phenotype data is collected from Chromosome 6 Project participants (parent cohort) and eligible case reports (literature cohort) using the Chromosome 6 Questionnaire (see Supplementary Information S1 for further details). This multilingual web-based questionnaire contains 132 main questions that cover 654 phenotypic features, developmental characteristics and other types of clinical information (Supplementary Table S1). All phenotypes are coded using Human Phenotype Ontology^8^ (*n* = 423), ICD-10^9^ (*n* = 11), ORPHA^10^ (*n* = 1) or custom (*n* = 219) terms. Custom codes were developed to encode information for which no standard codes were available, such as growth parameters, age at onset of certain conditions, or developmental milestones. Genotype data from both cohorts derive from the results of microarray or other high-resolution genetic analyses, performed in different laboratories using various platforms. Genomic locations based on a reference genome other than hg19 are converted to hg19 using the UCSC LiftOver Tool (https://genome.ucsc.edu/cgi-bin/hgLiftOver).^11^

Del2Phen annotates several aspects of each chromosome aberration, including type (deletion versus duplication), locus (base pair position at the start and end of the aberration in hg19) and genes intersecting the aberration. For each gene, the following haploinsufficiency (HI) scores are annotated: HI score from Huang et al. (2010)^12^, probability of loss-of-function intolerance (pLI) score based on Lek et al. (2016),^13^ and predicted probability of haploinsufficiency (pHaplo) score from Collins et al. (2022).^14^ Each score denotes the probability of a gene having a clinical effect in case of deletions or loss-of-function variants. Genes with an HI score between 0-10%, a pLI score ≥0.9 or a pHaplo score ≥0.86 are most likely to have a phenotypic effect.^12–14^ For Del2Phen, genes are considered HI-genes if they fulfil at least two of these three criteria when all three scores are known or at least one of two criteria when only two scores are known. Selected HI-genes known to have a prominent, highly penetrant effect on the phenotype are annotated as dominating-effect genes in the tool. Details on the selection of these genes and additional technical information are provided in Supplementary Information S1.

### 2.2. Workflow and genotypic similarity settings

Del2Phen performs pairwise comparisons between patients using specific parameters to score their genotypic similarity. For aberrations of the same type on the same chromosome, the Jaccard index is used to measure the size, location and (HI-)gene overlap between pairs (see example in Figure S1). For each of these parameters, minimum thresholds can be introduced. Dominating-effect gene content is the only binary parameter, indicating whether or not the sets of affected dominating- effect genes are equivalent (concordant) between patients. For patients harbouring multiple aberrations of the same type on the same chromosome, Del2Phen uses the total size and gene content of these aberrations for the comparison.

Given the prominent role that dominating-effect genes play in the clinical picture, Del2Phen’s first genotypic similarity parameter is concordance in the presence or absence of these genes. Only individuals with the same affected dominating-effect genes are identified as genotypically similar by the tool. Similarly, because of their predicted phenotypic effect, Del2Phen’s second genotypic similarity parameter is the fraction of HI-genes shared between the index and each individual concordant for the selected dominating-effect genes. This parameter, HI-gene similarity (Figure S1), can range from 0 to 100% depending on the extent of HI-gene overlap between deletions. Del2Phen uses these two parameters to identify individuals in the Chromosome 6 database as genotypically similar to an index, thereby generating a phenotype description tailored to that index patient (Figure 1).

### 2.3. Assessing optimal HI-gene similarity for chromosome 6 deletions

Ideally, Del2Phen would only generate a phenotype description based on individuals with 100% HI- gene overlap, ensuring the highest level of functional similarity. However, this would result in too few genotypically similar cases. To identify the thresholds that would allow as many individuals with a chromosome 6 deletion as possible to receive a phenotype description based on highly similar deletions, we evaluated the impact of various minimum HI-gene similarity thresholds on the: (i) size of the genotypically similar group (i.e. group size or number of connections) and (ii) precision of the resulting phenotype descriptions, quantified using positive predictive value (PPV). We used 115 main features to determine PPV (features in bold in Table S1) to prevent double-counting of the same feature if both the main and sub-features were known (see Supplementary Information S1 for additional criteria).

PPV was calculated individually for all patients in the database who had at least five connections for any given HI-gene similarity (each patient was used as an index once) using the following formula: PPV = *number of features in the description that are also present in index* / *total number of features in the description*, where ‘*total number of features in the description*’ represents the number of bold features in Table S1 that were present in ≥20% of the connected group *and* at least two individuals. The ‘*at least two individuals’* criterion was added to exclude features present in only one patient in case of a minimum number of connections (*n* = 5). For these calculations, features not reported in literature cases were considered not present.

The objective of our assessments was to determine the extent to which we can decrease the minimum HI-gene similarity in order to reach higher numbers of genotypically similar cases while still preserving PPV. Given the non-normal data distribution, we used the median (50^th^ quartile) as a measure of PPV. To ensure that the presence of a dominating-effect gene does not affect the overall results, this metric was also determined separately for individuals with and without such a gene in their deletion, and any significant differences between these groups were evaluated using a Mann-Whitney U test. As the sample sizes in the literature and parent cohorts were insufficient for separate assessments, we examined differences in the number of phenotypic features (Mann-Whitney U test) and dominating-effect gene content (Chi-square test) between the two cohorts to address potential confounding. All statistical analyses were performed with Python 3.10^15^ using the SciPy,^16^ Matplotlib^17^ and Plotly packages.^18^

Lastly, to demonstrate Del2Phen’s application using the genotypic similarity parameters determined in this study, we used it to generate a phenotype description for two randomly selected patients in the Chromosome 6 database: one with a dominating-effect gene in their deletion and one without.

## 3. Results

### 3.1. Participant and genotype characteristics

As of November 2023, the Chromosome 6 database included detailed genotype and phenotype information for 184 project participants who had completed the Chromosome 6 Questionnaire (parent cohort) and 342 individuals described in literature (literature cohort). From these, 452 individuals (142 from the parent cohort and 310 from the literature cohort) were eligible for use by Del2Phen (Table 1 and Figure S2). Their deletions were not equally distributed throughout the chromosome (Figure S3), with the proximal 6p region having a notably low number of cases. The number of phenotypic features per patient was significantly higher in the parent cohort compared to the literature cohort (*p* <.0001) (Table 1), but there was no significant difference between cohorts in the number of cases encompassing at least one dominating-effect gene (*p* =.28).

**Table 1.**
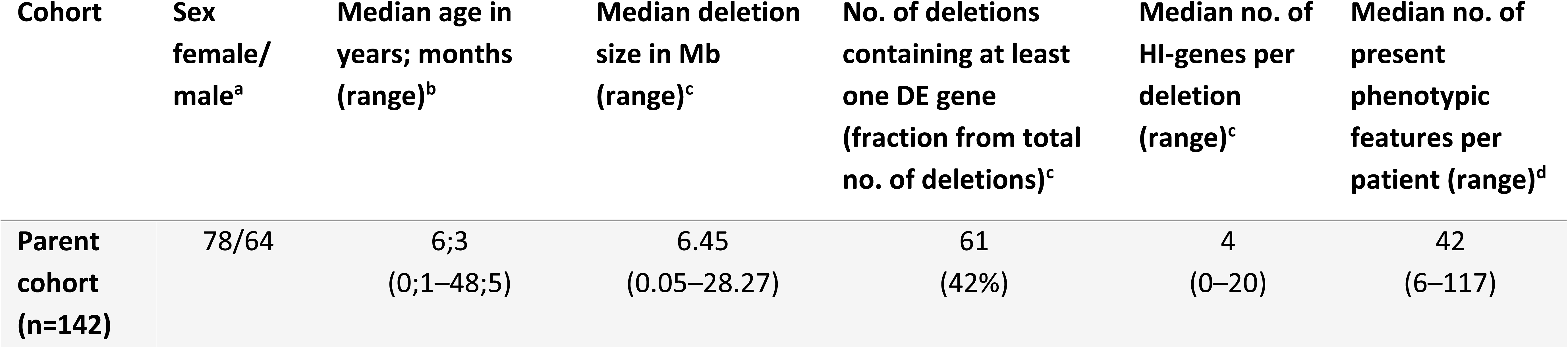
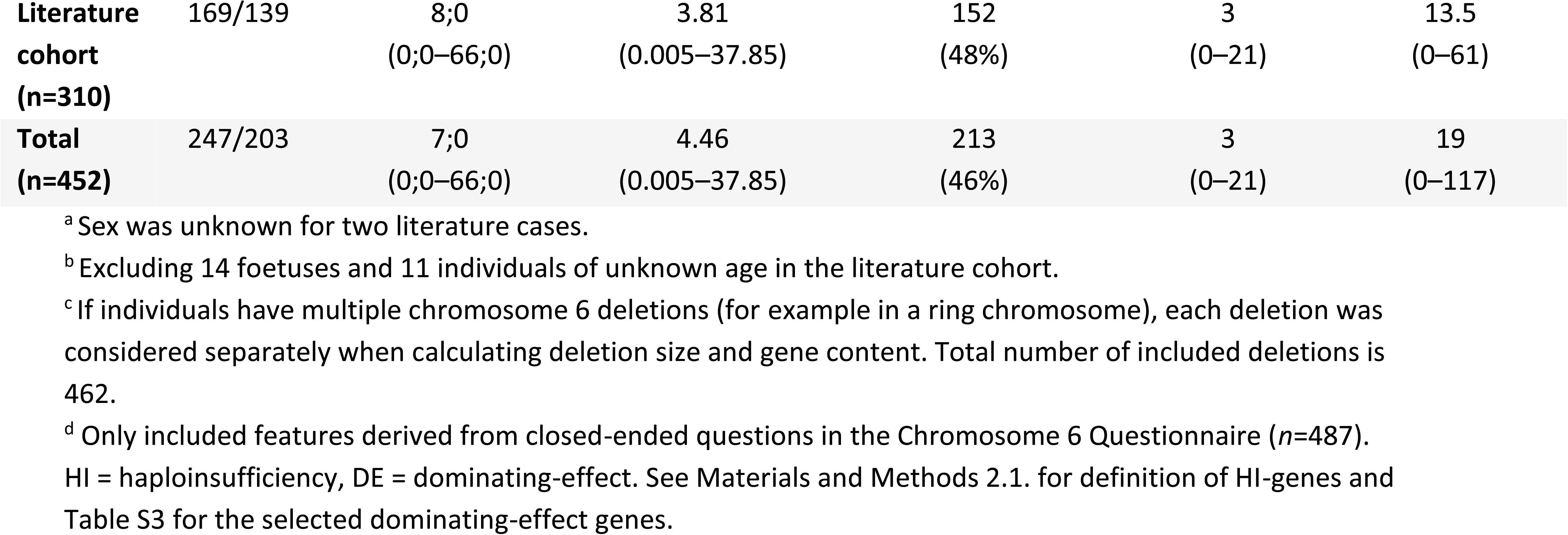
Characteristics of patients included in the current study.

| Cohort | Sex female/<br>male<sup>a</sup> | Median age in<br>years; months<br>(range)<sup>b</sup> | Median deletion<br>size in Mb<br>(range)<sup>c</sup> | No. of deletions<br>containing at least<br>one DE gene<br>(fraction from total<br>no. of deletions)<sup>c</sup> | Median no. of<br>HI-genes per<br>deletion<br>(range)<sup>c</sup> | Median no. of<br>present<br>phenotypic<br>features per<br>patient (range)<sup>d</sup> |
| --- | --- | --- | --- | --- | --- | --- |
| Parent<br>cohort<br>(n=142) | 78/64 | 6;3<br>(0;1–48;5) | 6.45<br>(0.05–28.27) | 61<br>(42%) | 4<br>(0–20) | 42<br>(6–117) |
| Literature cohort (n=310) | 169/139 | 8;0<br>(0;0–66;0) | 3.81<br>(0.005–37.85) | 152<br>(48%) | 3<br>(0–21) | 13.5<br>(0–61) |
| Total (n=452) | 247/203 | 7;0<br>(0;0–66;0) | 4.46<br>(0.005–37.85) | 213<br>(46%) | 3<br>(0–21) | 19<br>(0–117) |

<sup>a</sup> Sex was unknown for two literature cases.
<sup>b</sup> Excluding 14 fetuses and 11 individuals of unknown age in the literature cohort.
<sup>c</sup> If individuals have multiple chromosome 6 deletions (for example in a ring chromosome), each deletion was considered separately when calculating deletion size and gene content. Total number of included deletions is 462.
<sup>d</sup> Only included features derived from closed-ended questions in the Chromosome 6 Questionnaire (n=487).
HI = haploinsufficiency, DE = dominating-effect. See Materials and Methods 2.1. for definition of HI-genes and Table S3 for the selected dominating-effect genes.

### 3.2. Assessment of genotypic similarity parameters

Of the 121 genes (Table S2) that fulfilled our definition of a HI-gene (Materials and Methods 2.1.), five were selected as dominating-effect genes and used as the tool’s first genotypic similarity parameter (Table S3).

As expected, the number of individuals who exceed a certain number of connections (5, 15, 20 or 30) decreases as we increase the minimum HI-gene similarity threshold (Table 2 and Figure S4). Conversely, the overall median PPV, calculated across all eligible patients, shows a positive trend with HI-gene overlap, peaking at 0.333 when minimum HI-gene similarity is set to 70% or higher (Table 2). Interestingly, the increase in median PPV with HI-gene similarity is only marginal in groups containing a dominating-effect gene (Figure 2A), with PPV being significantly higher in these groups at lower HI-gene similarity thresholds (≤50%) when compared to groups without such a gene (Table S4). This difference is no longer statistically significant when the minimum HI-gene similarity is at least 60%.

**Fig. 2:**
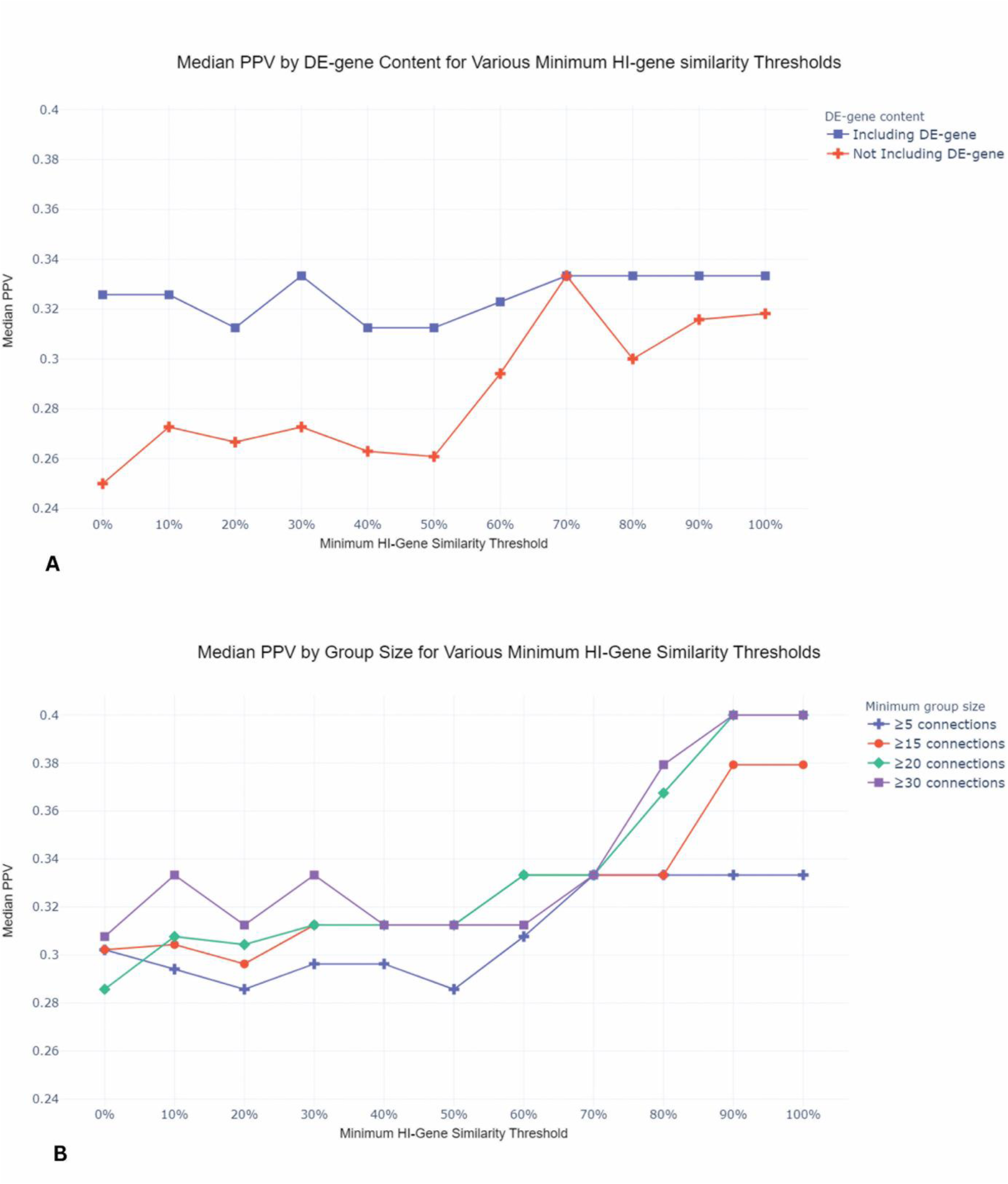
Median PPV based on dominating-effect (DE) gene content (A) and minimum group size (B) for various minimum HI-gene similarity thresholds. **A.** Median PPV for individuals harbouring a DE- gene in their deletion exhibits only slight variation in response to changes in the minimum HI-gene similarity threshold. In contrast, for individuals who do not lack such a gene, median PPV notably increases when the minimum HI-gene similarity threshold surpasses 50%. **B.** When comparing groups of different sizes, larger groups (≥15 or ≥20 individuals) tend to exhibit higher median PPVs, particularly at higher HI-gene similarities (≥80%). At these similarity thresholds, increasing the minimum group size from 20 to 30 does not affect PPV. For HI-gene similarities thresholds of ≥60% and ≥70%, groups of ≥15 and ≥20 individuals exhibit the same median PPV (0.333).

**Table 2.** Effect of increasing minimum HI-gene similarity threshold on group size and PPV.

| Minimum HI-gene similarity threshold | Number of individuals with $\geq 15/\geq 20/\geq 30$ connections | Median PPV overall <sup>a</sup> | Median PPV if $<15/\geq 15$ connections <sup>b</sup> | Median PPV if $<20/\geq 20$ connections <sup>b</sup> | Median PPV if $<30/\geq 30$ connections <sup>b</sup> |
| --- | --- | --- | --- | --- | --- |
| 50% | 251/215/156 | 0.286 | **0.231/0.313** | **0.25/0.313** | **0.267/0.313** |
| 60% | 205/165/84 | 0.308 | **0.265/0.333** | **0.273/0.333** | 0.304/0.313 |
| 70% | 161/108/81 | 0.333 | **0.297/0.333** | 0.333/0.333 | 0.333/0.333 |
| 80% | 148/108/81 | 0.333 | 0.316/0.333 | 0.313/0.368 | 0.333/0.379 |
| 90% | 103/85/35 | 0.333 | **0.3/0.379** | **0.3/0.4** | 0.32/0.4 |
| 100% | 103/85/35 | 0.333 | 0.3/0.379 | **0.3/0.4** | 0.327/0.4 |

<sup>a</sup> Overall values indicate the PPV as determined for all individuals for whom this metric could be calculated (individuals with at least 5 connections), irrespective of group size.
<sup>b</sup> Bold values indicate a statistically significant difference ( $p < 0.5$ ). $p$ -values can be found in Table S5.
HI = haploinsufficiency, PPV = positive predictive value. See Materials and Methods 2.1. for definition of HI-genes, and section 2.3.1. for definition and calculation of PPV. Dominating-effect gene content was always taken into account when grouping.

When evaluating groups based on different minimum sizes, the highest median PPV (0.4) occurs in individuals with ≥20 connections at HI-gene similarity thresholds of ≥90% and 100%, and increasing the group size to ≥30 does not further improve this metric (Table 2 and Figure 2B). When the minimum HI-gene similarity is <80%, the differences between groups of different sizes become less pronounced. At ≥60% and ≥70% HI-gene similarity, groups containing ≥15 individuals and those with ≥20 exhibit a similar median PPV (0.333) (Figure 2B), with a minimum group size of 15 individuals sufficient to achieve a statistically significant increase in PPV compared to smaller groups at these thresholds (*p*-values provided in Table S5).

### 3.3. Del2Phen’s default genotypic similarity settings for chromosome 6 deletions

Considering the results above, we determined the genotypic similarity algorithm depicted in Figure 3 to guide Del2Phen’s generation of phenotype descriptions for chromosome 6 deletions. Under the default settings, at HI-gene similarity ≥80%, Del2Phen will provide a clinical description with the highest achievable functional similarity for groups of ≥20 individuals. At 60% and 70% HI-gene similarity, the group-size cutoff will be reduced to 15. Below these similarity thresholds, phenotype descriptions will only be generated if a dominating-effect gene is present, coupled with the highest HI-gene similarity that would yield groups of at least 15. The rationale behind this algorithm is discussed in section 4.3.

**Fig. 3:**
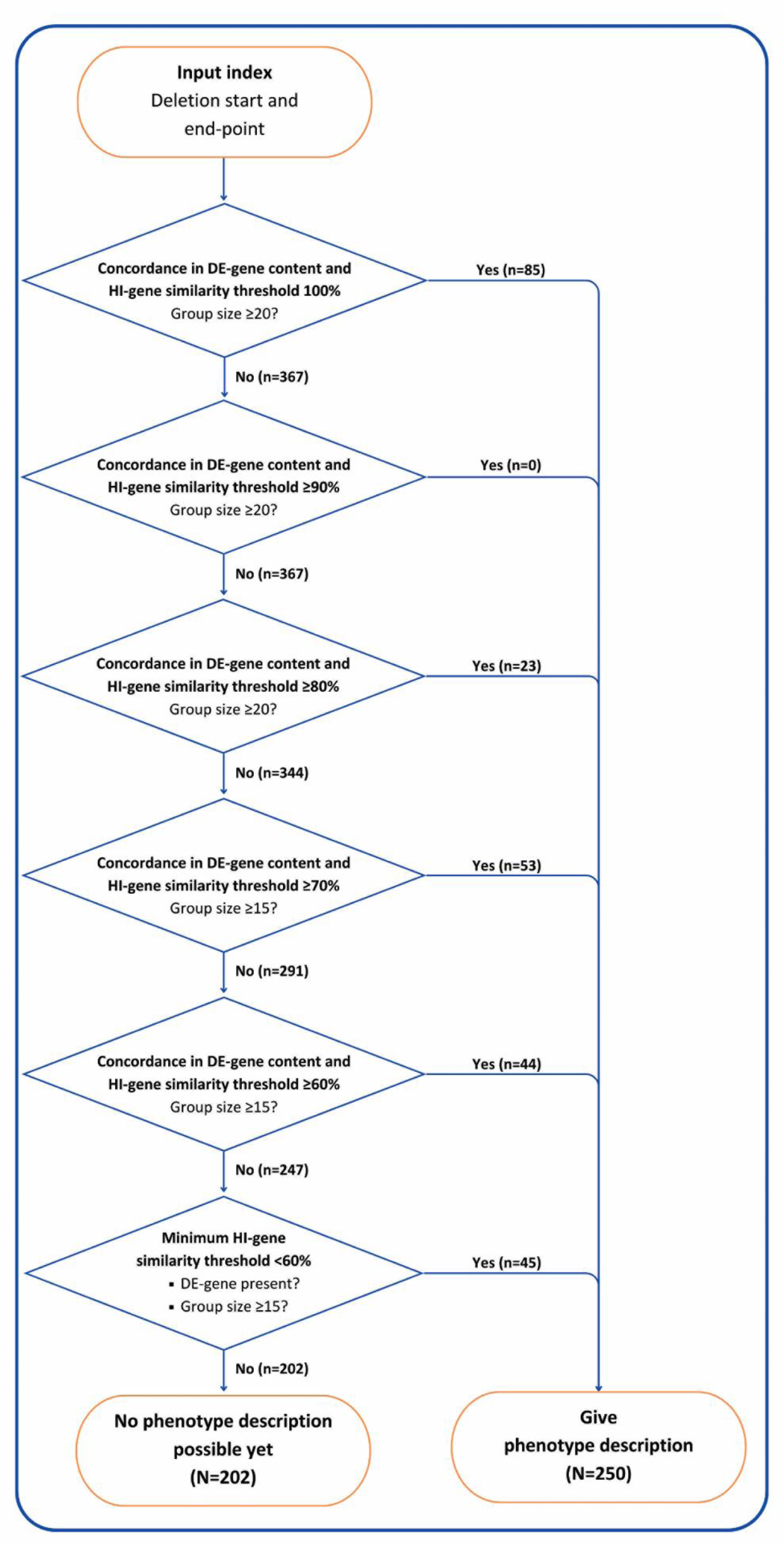
Genotypic similarity algorithm for Del2Phen’s generation of phenotype descriptions for chromosome 6 deletions. The tool will leverage the maximum possible functional similarity within groups that exceed a certain size for generating a comprehensive phenotype description. At HI-gene similarity ≥80%, a description will be provided for groups of at least 20 individuals. The group size cutoff will be lowered to 15 for HI-gene similarities of at least 70% and 60%. Below these similarity thresholds, phenotype descriptions will only be given if a dominating-effect (DE) gene is present, coupled with the highest HI-gene similarity yielding groups of at least 15 individuals. In each step, *n* represents the number of additional individuals in the Chromosome 6 database who become eligible for a phenotype description as the algorithm progresses through the step, while *N* represents the total number of individuals who would or would not receive a phenotype description based on these settings and our current data pool.

Table 3 and Table S6 show the phenotype descriptions for two patients from the Chromosome 6 database generated by Del2Phen based on this algorithm. The first patient (individual Id226) has a 2.4 Mb 6q24.3q25.1 deletion that includes the dominating-effect gene *TAB2* (TAK1- BINDING PROTEIN 2, HGNC:17075, MIM* 605101). Del2Phen identified 17 individuals in the Chromosome 6 database (2 Chromosome 6 Project participants and 15 literature cases) who shared the same dominating-effect gene and at least 70% of their HI-gene content with this patient. The phenotype description for this patient (Table 3) consists of 25 features present in at least 20% of the connected group. Of these, 12 features were also seen in the index, leading to a PPV of 0.48 based on the full description and a PPV of 0.6 when considering only the selected features used to determine the tool’s setting (Table S1).

**Table 3.** Example of a clinical description generated by Del2Phen for a patient with a dominating- effect gene in the deletion.

| Clinical description for patient Id226 | Features present in patient Id226 |
| --- | --- |
| **Intrauterine growth retardation** | **Intrauterine growth retardation** |
| Hospitalisation after birth | Hospitalisation after birth |
|  | *Hospitalisation due to congenital problems* |
|  | Premature rupture of membranes |
|  | *Premature birth* |
|  | **Neonatal asphyxia** |
|  | **Prolonged neonatal jaundice** |
|  | *Feeding problems in infancy* |
| **Abnormality of the outer ear** |  |
|  | **Abnormality of the skin** |
|  | Hypopigmented skin patches |
|  | Capillary haemangioma |
| **Joint hypermobility** | **Joint hypermobility** |
| Generalised joint laxity | Generalised joint laxity |
| **Abnormal hand morphology** |  |
|  | **Abdominal wall defect** |
|  | *Umbilical hernia* |
| **Sacral dimple** | **Sacral dimple** |
| Abnormality of body height | Abnormality of body height |
| **Short stature** | **Short stature** |
| **Abnormality of the respiratory system** |  |
| **Abnormal heart morphology** | **Abnormal heart morphology** |
| Abnormal cardiac septum morphology | Abnormal cardiac septum morphology |
| Abnormal atrial septum morphology |  |
| Abnormal heart valve morphology | Abnormal heart valve morphology |
| Abnormal aortic valve morphology |  |
| Bicuspid aortic valve |  |
| Abnormal tricuspid valve morphology | Abnormal tricuspid valve morphology |
| Abnormal mitral valve morphology |  |
| Patent ductus arteriosus |  |
| **Cardiomyopathy** |  |
| Dilated cardiomyopathy |  |
| **Muscular hypotonia** | **Muscular hypotonia** |
|  | **Seizure** |
|  | Febrile seizure |
|  | *Normal hearing at newborn screening* |
| Neurodevelopmental delay |  |
| Delayed gross motor development |  |
| Delayed speech and language development |  |

|  | *Social behaviour* |
| --- | --- |
|  | *Helpful behaviour* |
| **Phenotype description assessment** |  |
| **Total number of features in description** | 25 |
| **(only using the selected feature in bold)** | (10) |
| **Number of features in description seen in patient** | 12 |
| **(only using the selected feature in bold)** | (6) |
| **PPV based on full description** | 0.48 |
| **(only using the selected feature in bold)** | (0.6) |

The second phenotype description (Table S6) was generated for a patient with an 11.1 Mb deletion in the 6q13q14.1 region that does not span a dominating-effect gene. The description is based on the phenotypes of 16 individuals (5 project participants and 11 literature cases) with a minimum of 70% overlap in their HI-gene content. The clinical description for this patient was lengthier, encompassing 38 features, and resulted in a PPV of 0.45, rising to 0.5 when using only the selected features in Table S1.

The clinical description for this patient is based on the phenotype and other clinical features of 17 individuals that share the same content of dominating-effect genes and at least 70% of the affected HI-genes. The features in the description are present in at least 20% of the individuals connected to this patient (i.e. at least 4). The clinical description in this table is generated based on all features covered by the Chromosome 6 Questionnaire deriving from closed-ended questions (*n*=487). The features in bold were used to calculate PPV in this study. In italics are the features of the index that were also seen in at least one connected individual but did not meet the prevalence threshold for inclusion in the description (i.e. were seen in less than 20% of the connected group).

## 4. Discussion

We have introduced Del2Phen, an innovative tool developed to provide tailored phenotype descriptions for chromosome aberrations. In this study, Del2Phen was applied to deletions on chromosome 6 based on the data collected through the Chromosome 6 Project. We have described the process through which Del2Phen’s genotypic similarity parameters were evaluated, with the aim of determining its definitive settings for chromosome 6 deletions. Below, we discuss this optimisation process, the rationale behind Del2Phen’s genotypic similarity algorithm, and the future steps for this tool to be implemented in practice.

### 4.1. Phenotype descriptions for chromosome deletions

Defining the functional similarity of chromosome aberrations is complex because of our still incomplete understanding of the function of many genes and the clinical consequences in case of deletions. While websites aimed at professionals, such as DECIPHER,^19^ often allow users to set the functional similarity range, Del2Phen will eventually be incorporated into a website primarily for parents. It is therefore important to pre-set the genotypic similarity parameters that will be used to generate phenotype descriptions. For Del2Phen, we utilised current knowledge of the predicted HI-effect of genes and known gene-phenotype associations to define genotypic similarity.

Because of the dominating role of the genes in Table S3 in the clinical picture, we expect individuals harbouring identical sets of these genes to have similar phenotypes. The observed higher PPV in groups harbouring a dominating-effect gene even at otherwise low HI-gene similarity (Figure 2A and Table S4), confirms the significant role of these genes in the clinical outcome and supports our decision to use concordance in dominating-effect gene content as Del2Phen’s first genotypic similarity parameter. This is also illustrated by the example in Table 3, where all the features associated with a deletion of the dominating-effect gene *TAB2* (Table S3) were included in the phenotype description, and most were present in the index.

The second parameter, the HI-gene overlap between deletions, would ideally be set to 100%. However, the limited number of individuals who have fully overlapping deletions means that such high similarity would often result in phenotype descriptions based on small patient groups, if generated at all. Given the role of individual variation in the resulting phenotype and the incomplete penetrance of most observed features,^20^ it was necessary to lower the HI-gene similarity cutoff to enable a larger number of individuals to receive a phenotype description and increase the sample size on which these descriptions would be based. Concurrently, it was important to ensure that adjusting the HI-gene similarity would not compromise the reliability of Del2Phen’s outcome.

### 4.2. Evaluating phenotype descriptions

Assessing the accuracy of phenotype descriptions in the context of structural chromosome aberrations is inherently complex, as even identical deletions can result in an array of phenotypic manifestations.^21,22^ To gain insights into the relevance of features in the descriptions, we used PPV as an outcome measure to describe the fraction of features in the clinical description that were in accordance with the phenotype of the index. Remarkably, PPV did not drop below 0.2 when setting the HI-gene similarity threshold to 0% (Figure 2). While this may seem counterintuitive, these thresholds only indicate a minimum level of similarity, e.g. at a threshold of 10%, groups are formed of individuals who share 10% *or more* of their deletion’s HI-gene content. Increasing the minimum threshold results in individuals with lower levels of similarity being excluded from the groups, only retaining those sharing a greater fraction of HI-genes. This, in turn, contributes to a higher description PPV as the minimum HI-gene similarity increases, reaching a median of 0.333 for functionally identical deletions (Figure 2).

A maximum median PPV of 0.333 may seem low, but we have to consider the low *a priori* chance of multiple features occurring together in a single patient (index) given the reduced penetrance of many of the observed features and the variable expressivity associated with these aberrations.^20,21^ For instance, if a characteristic is present in 25% of the genotypically similar group, it will be included in the phenotype description under our current settings but may not be present in the index patient, thus decreasing PPV. We believe that it is correct to include these features in the description to ensure comprehensiveness and not miss important findings, even though this might lower the tool’s precision metrics. Furthermore, the decrease in group size at such high HI-gene similarity (Figure S4) can also influence PPV.

Another potential factor is the variability in phenotypes associated with deletions in certain chromosome regions. While deletions in certain areas are linked to highly distinct phenotypes,^4^ deletions in other regions produce a much more diverse clinical outcome,^5^ thereby affecting the overall PPV. However, as Del2Phen’s similarity settings will be applied uniformly across the entire chromosome, PPV was determined for deletions in all regions.

Lastly, our relatively modest PPV might also be influenced by the amount and diversity of phenotypic information available for our parent cohort. In contrast to literature reports, in which authors are often restricted in the amount and kind of information they present (e.g. presenting only clinically significant issues), data collected via the Chromosome 6 Questionnaire yields much more detailed information (Table 1).^7^ Since almost all groups consist of both literature cases and Chromosome 6 Project participants, one subset of the group will have fewer but very specific characteristics (literature cohort), while the rest will present with a much more diverse phenotype (parent cohort), thus lowering PPV. Unfortunately, it was not possible to conduct the analysis separately for the two cohorts. Nevertheless, we can conclude that the extensive information collected from our parent cohort makes the phenotype descriptions generated by Del2Phen more complete than what can be derived solely from literature reviews.

### 4.3. Determining Del2Phen’s genotypic similarity settings for chromosome 6 deletions

Following Del2Phen’s genotypic similarity algorithm (Figure 3), a phenotype description will be generated based on groups of ≥20 individuals sharing the highest attainable HI-gene fraction in the range of 80% to 100%. This minimum group-size threshold is justified by our observations that PPV is higher in groups of ≥20, with no notable improvements when increasing group size to ≥30 (Table 2 and Figure 2B). At the minimum HI-gene similarity thresholds 60% and 70%, the median PPV remained constant when lowering the group size to ≥15 individuals (Table 2), while still being significantly higher compared to groups <15 (Table S5). For this reason, the group-size cutoff was lowered to at least 15 at these similarity thresholds, allowing Del2Phen to provide phenotype descriptions for a larger fraction of individuals in the database, as well as future new patients (Figure S4).

If an insufficient number of patients in the database share ≥60% of their HI-genes, Del2Phen will still provide a description for deletions that include a dominating-effect gene, based on the highest possible HI-gene overlap that would yield groups of ≥15 (Figure 3). This setting is based on the expectation that the phenotype in these groups would be primarily determined by the deletion of the dominating-effect gene rather than by the HI-gene similarity, as supported by our findings in Figure 2A and Table S4. At HI-gene similarity ≥60%, the difference in PPV between groups with and without a dominating-effect gene is no longer significant, suggesting that this fraction of shared HI-genes is sufficient to produce effects similar in impact to that of a dominating-effect gene.

Based on these settings and our current data pool, 250 individuals (55% of our current cohort) will be eligible to receive an automatically generated phenotype description (Figure 3). As the Chromosome 6 database continues to grow, the number of individuals eligible for a phenotype prediction will also increase.

### 4.4. Strengths and limitations

Del2Phen is an innovative phenotype description tool, developed primarily for delivering clinical information to parents. Unlike existing online tools, which are tailored to professionals and require a certain level of expertise, Del2Phen will deliver information via a parent-friendly website. Naturally, it can also be a valuable resource for healthcare professionals in counselling patients and their families. The clinical descriptions provided will be detailed and thorough compared to the limited information in current databases or literature reports, because Del2Phen uses the extensive well-curated data collected via the Chromosome 6 Project (Table 1).^7^ Even though it is currently optimised for chromosome 6, it can also be applied to other chromosomes if given the correct input, and could thus benefit people affected by a wider spectrum of chromosome disorders.

Since Del2Phen will be periodically updated with the latest information on HI or the clinical effect of genes, the clinical information it produces will be more up-to-date than that in scientific articles, which reflect knowledge at the time of writing. Although Del2Phen does not specifically account for non-coding features, which are often positioned on fixed locations on the genome, their effects will be incorporated once more evidence is available. Lastly, this tool can also be applied in research settings to expedite data analysis.

One limitation of Del2Phen pertains to the fact that its output can be sensitive to variability in resolution of the genotype data, which derive mostly from microarray analysis conducted using different platforms. We do not expect this to affect the outcome of this study, as only individuals for whom a high-resolution array was available were included in the Chromosome 6 Project.

Furthermore, an HI-gene located between the last absent and first present probes (or *vice versa*) was an exclusion criterion for the current study as it made the deletion of that gene unclear (Supplementary Information S1), thus ensuring that the grouping of genotypically similar individuals would not be affected.

Clinical data missing from literature reports might have an impact on the thoroughness of the phenotype descriptions. To avoid falsely increasing the prevalence of certain features, we considered features that were not mentioned in the literature reports as not present in the patients. We expect this assumption to hold true for highly clinically relevant features, but articles that focus on specific topics, like heart or brain defects, may sometimes omit unrelated phenotypes.

Finally, it is important to note that we must always be cautious when presenting clinical information on chromosome aberrations, whether derived from automatic or manual analysis. The unique nature of these aberrations, paired with the phenotypic variability, means that not all features will be seen in all patients. Such disclaimers will be added to our website, together with instructions for reading the clinical descriptions, links to websites with support information, and the option to download the medical information for discussion with healthcare professionals.

### 4.5. Future steps

The next step for Del2Phen is validating its output by comparing its outcomes to those from manual data analysis, the conventional approach for characterising the phenotypes of chromosome deletions. Following this, the tool will be deployed to describe the phenotypes of new, uncharacterised regions. In parallel, the development of the interactive website for parents will also commence. In a running study, with the help of involved parents and professionals, we are investigating the best way to report the clinical features on the website. The study’s findings will inform the website’s content and structure, including the minimum prevalence of clinical features in individuals with a similar aberration required for inclusion in the description. Lastly, while Del2Phen currently focuses solely on deletions, it will be modified for use on duplications once enough data on these aberrations is collected.

## 5. Conclusion

We have introduced Del2Phen, a novel phenotype description tool for chromosome aberration. Descriptions are based on features present in individuals identified as genotypically similar by the tool. Genotypic similarity parameters were determined by leveraging existing knowledge on gene-phenotype relationships and the HI-effect of genes. Furthermore, we described the optimization process for this tool to produce thorough clinical descriptions for the full range of chromosome 6 deletions using data collected through the Chromosome 6 Project. Del2Phen will be incorporated into a website aimed at parents of affected children, providing them with essential health information in an efficient and timely manner and thus bridging the knowledge gap surrounding these rare disorders.

## Supporting information

README file for Del2Phen

## 6. Declaration of AI and AI-assisted technologies in the writing process

During the preparation of this work the authors used Open AI’s ChatGPT 4.0 in order to improve language and readability. After using this service, the authors reviewed and edited the content as needed and take full responsibility for the content of the publication.

## 7. Data Availability Statement

The phenotype and genotype data of Chromosome 6 Project participants are stored in the secure in-house Chromosome 6 database. The data is also available open-access on DECIPHER (www.deciphergenomics.org)19 under the following patient IDs: 425375, 425379-425388, 426261, 433748-433760, 489709-489746 and 524537-524630.

## 8. Code Availability

The code for Del2Phen is available on GitHub (https://github.com/Chromosome-6-Project/Del2Phen) and PyPI (pypi.org/project/del2phen). More details about the code and using the tool can be found in the supplementary README file.

## 9. Acknowledgments

We extend our gratitude to all participating patients and their families for contributing their data and making this research possible. We would like to thank Pauline Bouman in particular for her essential role as our contact parent for the Chromosome 6 Facebook support group. Our appreciation also goes to Kate Mc Intyre for editing this manuscript and to Imke Christiaans for her insightful ideas on data presentation and additional analyses. Lastly, we would like to thank the MOLGENIS team at the Genomics Coordination Center Groningen, especially Fernanda de Andrade, for the technical support with the Chromosome 6 database.

## 10. Funding

ER and AE are recipients of a JSM MD/PhD scholarship from the University Medical Center Groningen, part of which has been used for this study. This research was additionally supported by a grant from ZonMw (113312101) and crowdfunding organised by Chromosome 6 parents. MAS is recipient of a VIDI grant (917.164.455) from the Netherland’s Organisation for Scientific Research (NWO).

## 11. Author Contribution Statement

Conceptualisation: C.M.A.R.A., A.E.; Methodology: C.M.A.R.A., L.F.J., E.R., T.D.M. and A.E.; Software: T.D.M., M.K.S. and M.A.S.; Data curation: E.R. and T.D.; Formal analysis: T.D.M. and E.R.; Writing – original draft: E.R.; Writing – review and editing: T.D.M., C.M.A.R.A., M.K.S., L.F.J., A.E., T.D. and M.A.S.; Visualisation: E.R. and T.D.M.; Supervision: C.M.A.R.A., L.F.J. and A.E.; Funding acquisition: E.R., A.E., C.M.A.R.A. and M.A.S. The final version of this manuscript has been approved by all authors.

## 12. Ethics Declaration

A complete ethical evaluation was waived by the accredited Medical Ethics Review Committee of the University Medical Center Groningen. According to Dutch guidelines, research conducted anonymously with existing data without performing additional investigations does not fall under scientific research as described in the Medical Research Involving Human Subjects Act (WMO), and therefore does not require ethical approval.

Informed consent for participation in the study and publication of the results was obtained from all participants as part of signing up for the Chromosome 6 Project. The project’s privacy statement is available on the website: chromosome6.org/privacy-statement

## 13. Conflicts of interest

The authors declare no conflicts of interest.

## 14. Supplementary Material

The Supplementary Information file includes the supplementary methods, Figures S1-S4 and Tables S3-S6. Tables S1 and S2 and provided as separate Excel files. The supplementary README file provides additional technical details on Del2Phen as well as instructions on how to use it.

## Notes

### Competing Interest Statement

The authors have declared no competing interest.

### Clinical Protocols

https://github.com/Chromosome-6-Project/Del2Phen

### Funding Statement

ER and AE are recipients of a JSM MD/PhD scholarship from the University Medical Center Groningen. This research was additionally supported by a grant from ZonMw (113312101) and crowdfunding organised by Chromosome 6 parents. MAS is recipient of a VIDI grant (917.164.455) from the Netherlands Organisation for Scientific Research (NWO).

### Author Declarations

The accredited Medical Ethics Review Committee of the University Medical Center Groningen waived ethical approval for this work.

