## Supplementary material for "Developing Del2Phen: a novel phenotype description tool for chromosome deletions": README file for Del2Phen

### README: Del2Phen

#### Introduction

Del2Phen is a tool for predicting phenotypes for a given copy number variant (CNV), based on known phenotypes from patients with similar CNVs. Patients are first grouped according to minimum genotypic similarity thresholds across multiple CNV properties, including genes and haploinsufficient genes affected by the CNV. Phenotypes are then predicted by the prevalence of phenotypes found in individuals within the group.

This project is part of [The Chromosome 6 Project], a research initiative that aims to provide information on rare chromosome 6 aberrations in affected children to parents and healthcare professionals.

#### Installation

Del2Phen is written in Python 3 and requires Python 3.9 or higher (except Python 3.10). We recommend using `conda` to create a Python 3.12 environment, then installing with `pip`:

```
conda create -n del2phen_env python==3.12
conda activate del2phen_env
python -m pip install del2phen
```

#### Getting Started

At minimum, Del2Phen requires basic patient CNV and phenotype tabular data.

##### CNV data

CNV input data must be provided as a 5-column tab-separated file. Each row should specify one CNV from one patient. Patients with multiple CNVs should be listed with the same ID on multiple lines, one CNV per line. The file must include the following headings:

- `id`: Patient ID of the CNV. The same ID can be used multiple times to indicate that one patient has multiple CNVs.
- `chromosome`: Chromosome on which the CNV is located. These chromosome names must match the names used in the geneset GTF file used (e.g., make sure that your GTF file uses either 'chrX' or 'X' for the chromosome names, consistently).
- `start`: 1-indexed inclusive CNV start position.
- `stop`: 1-indexed inclusive CNV stop position.
- `copy_number`: Integer value of the CNV, e.g., a single copy deletion is 1, a single copy duplication is 3.

##### Phenotype data

Patient phenotype data must be provided as a tab-separated file, in which the first column contains patient IDs with the header `id`. Each additional column must have as a header either a [Human Phenotype Ontology (HPO)] phenotype ID (e.g., `HP:0001643`) or any other identifier, which must then be defined as a [custom phenotype] term. Each entry in each column should be coercible to a Boolean or NA (t, T, true, True, f, F, false, False, 0, 1, NA, or an empty value) representing whether or not the patient exhibits the phenotype. Patients are not required to be present in the phenotype file to be present in the CNV file.

**Custom phenotypes**

Custom phenotypes (i.e., non-HPO identifiers) in the phenotype data must be defined in a separate tab-separated file with two columns with the headers `term_id` and `label`. `term_id` will be used as the unique identifier per phenotype, and `label` will be used as a display label.
See del2phen/resources/custom_phenotypes.tsv as an example.

#### Prediction

There are several ways to produce phenotype predictions with Del2Phen. The simplest way to predict phenotypes for a single query CNV is the following:

```
del2phen -g cnvs.tsv -p phenotypes.tsv -cnv chr6:123456-234567:1 --hi-gene-sim
0.75 -x -op ./
```

The above command will produce a table of phenotypes predicted for patients with a deletion (copy number of 1) on chromosome 6 from bases 123456 to 234567, and it will base the phenotype description only on patients who have at least 75% overlap in affected haploinsufficient genes. No other predictions for patients in the `cnvs.tsv` file will be produced due to using `-x / --cnv-query-only`, and results for the queried CNV will be written in `cnv_query_predictions.tsv` in the directory specified in `-op / --output-predictions`. Note that without specifying `-op / --output-predictions`, no predictions will be made.

Phenotypes can be predicted for one hypothetical individual with multiple CNVs by using the `-cnv / --cnv-predict` flag multiple times:

```
del2phen -g cnvs.tsv -p phenotypes.tsv -cnv chr6:123456-234567:1 -cnv
chrX:1357911-2468101:1 --hi-gene-sim 0.75 -x -op ./
```

To predict phenotypes for multiple individuals with different CNVs, add additional entries for these individuals to your `cnvs.tsv` file with a unique identifier for each individual. In this case, do not use the `-x / --cnv-query-only` flag and instead use the `--keep-unphenotyped` flag to predict phenotypes for these individuals:

```
del2phen -g cnvs.tsv -p phenotypes.tsv --keep-unphenotyped --hi-gene-sim 0.75 -op
./
```

If `-x / --cnv-query-only` is not specified but `-op / --output-predictions` is specified, predictions will be made for *all* patients in `-g/--genotypes` even if they already have phenotypes described in `-p/--phenotypes`. By specifying `-os / --output-stats` with an output file, precision metrics comparing the predicted phenotypes to the known phenotypes in `-p/--phenotypes` will be written per patient in a single table. This is useful for establishing effective comparison thresholds. You can omit `-op / --output-`

`predictions` to avoid excessive prediction tables while still outputting prediction metrics for testing:

```
del2phen -g cnvs.tsv -p phenotypes.tsv --hi-gene-sim 0.75 -os
prediction_metrics.tsv
```

#### Configuration

##### Filtering settings

There are several filtering options for subsetting which patients can be used for phenotype prediction to allow flexibility in analysis without the need to directly edit the `-g/--genotypes` and `-p/--phenotypes` TSV files.

- A text file with one patient ID per line can be provided with `-d / --drop-list` to specify patients to completely ignore.
- By default, Del2Phen will consider all CNVs per patient when comparing to the queried patient/CNV(s). For example, querying `-cnv chr2:100-200:1 --loci-similarity 0.8` will match a patient in `-g/--genotypes` with a CNV chr2:100-200:1, as they overlap 100%. However, this will *not* match a patient who has *both* a CNV chr2:100-200:1 and a CNV chr3:400-500:1, as they share only 50% of their total potentially-shared loci, which falls below the specified threshold of 80%. Comparisons can be restricted to consider only specific chromosomes/contigs by specifying `-c / --included-contigs`, followed by one or more contig names, space-separated.
- Similarly, analysis can be restricted to only consider specific copy number CNVs using `-cn / --included-copy-numbers`, followed by one or more copy number integers, space-separated, so that deletions are only compared with deletions and duplications only with duplications.

##### Genotype similarity settings

There are five criteria available for comparing patients to one another based on their genotypic similarity, i.e., how similar their CNVs are. If a comparison patient meets *all* the specified criteria when compared to the queried patient/CNV(s), then the comparison patient can be used to predict phenotypes.

- Length: Requires a minimum overlap in the combined total length of CNVs for patients to be compared using `--length-sim`. This is calculated as the Jaccard index between the total CNV lengths of each pair of patients being compared. Default=0, range=[0, 1].
- Loci: Requires a minimum overlap in the combined loci of CNVs for patients to be compared using `--loci-sim`. This is calculated as the Jaccard index between the

total sets of affected loci for each pair of patients being compared. Default=0, range=[0, 1].

- Affected genes: Requires a minimum overlap in the genes affected by the CNV of each patient using `--gene-sim`. This is calculated as the Jaccard index between each patient's set of CNV-affected genes. Default=0, range=[0, 1].

- Affected haploinsufficient genes: Requires a minimum overlap in the predicted haploinsufficient genes affected by the CNV of each patient using `--hi-gene-sim`. This is calculated as the Jaccard index between each patient's set of CNV-affected haploinsufficient genes. Default=0, range=[0, 1].

- Affected dominating-effect genes: Users can specify genes to be considered differently from other genes using `-de / --dominant-effect-genes` with a space-separated list of gene IDs, or using `--de-file` to specify a text file containing gene IDs, one per line. Specifying `--de-file` will override `-de / --dominant-effect-genes` if that is also present. By default, a comparison patient can only be used to make predictions if they share the same exact set of affected specified dominating-effect genes with the query patient/CNV(s) (note: if neither the query nor the comparison patient have *any* of these genes affected, they are considered a match and satisfy this criteria). Using the `--allow-de-gene-mismatch` flag disables this criterium. Alternatively, not specifying any of these genes achieves the same effect. See DE genes and References [1–7] for more information on the selected dominating-effect genes for chromosome 6.

#### Gene settings

##### HI genes

Del2Phen includes 3 haploinsufficiency (HI) prediction metrics from published studies: HI score, pHaplo score, and pLI score [8–10]. By default, genes in the provided gene set are considered haploinsufficient if they meet the default criteria of haploinsufficiency from at least 2/3 metrics if all 3 metrics are available for a gene, or only metric if 2 metrics are available. The default thresholds of each HI metric are taken from their original studies; however, these thresholds can be altered at runtime by using `-pli / --pli-threshold`, `-hi / --hi-threshold`, and `-phaplo / --phaplo-threshold`. The requirement for how many metrics must agree or how many metrics must be available for a gene to be considered haploinsufficient can be altered using `-m / --hi-mode` with the following options:

- `confirm`: Default behaviour as defined above.

- `any`: Only requires that any number of metrics must be available and any number of metrics must satisfy the threshold for haploinsufficiency.

- `2`: Require that at least 2 metrics must be available and that two satisfy their thresholds for haploinsufficiency.
- `all`: Require that all 3 metrics must be available and that all meet their thresholds for haploinsufficiency.

**DE genes**

Del2Phen can group patients based on the presence of genes that produce highly penetrant phenotypes when affected by a CNV (called "dominating-effect genes", or DE genes, here). By default, Del2Phen includes an example list of several such DE genes on chromosome 6 (see References [1–7]). To provide your own list of these genes, use `-de / --dominant-effect-genes` to list their gene IDs on the command line, or `--de-file` to provide a file listing them. To prevent any grouping by DE genes, use `--allow-de-gene-mismatch`.

#### Phenotype prediction thresholds

Once a group of patients satisfying the requirements of the genotype similarity settings is identified, phenotypes from this group are only predicted if they also meet minimum prevalence thresholds, which can be set with the following options:

- An absolute *minimum* number of patients in the group that exhibit the phenotype can be specified with `-abs / --absolute-threshold` (default=2).
- A relative *minimum* proportion of patients in the group that exhibit the phenotype can be specified with `-rel / --relative-threshold` (default=0.2).
    - This proportion is calculated, by default, as the number of patients that exhibit the phenotype ("True" response) out of all identified patients ("True", "False", *and* "NA" responses). This behaviour can be modified with `--ignore-nas` to ignore patients with an NA response, such that the proportion is calculated as True out of True + False responses only, within the identified patient group.
- A minimum group size can be required, such that if an insufficient number of genotypically similar patients are discovered, no phenotypes are reported, using `--group-size`.

#### Reference gene information

Del2Phen requires reference genome information to compare the gene content of CNVs between patients. This information includes the location of each gene and haploinsufficiency prediction scores. Del2Phen is packaged with 4 reference files in del2phen/resources/ and uses these by default:

- **hg19.ensGene.transcripts.gtf.gz**: Gene locus information from Ensembl for human genome **build 19** (hg19)
- **gnomad.v2.1.1.lof_metrics.by_gene.tsv**: pLI haploinsufficiency scores for hg19

- **HI_Predictions.v3.bed**: HI haploinsufficiency scores for hg19
- **phaplo.tsv**: pHaplo haploinsufficiency scores for hg19

Other reference files can be provided using `--gtf-file`, `--pli-file`, `--hi-file`, and `--phaplo-file`. Ensure that the gene IDs and genome builds are consistent.

**NOTE**: To prevent inadvertent mixing of genome builds/reference files, using any of the above arguments will stop Del2Phen from using *any* of the pre-packaged default files. **If, for example, you supply your own updated pHaplo score file, you *must* also provide, at minimum, a GTF file. Analysis will proceed without the other two haploinsufficiency score files, but no pLI or HI metrics will then be included in the analysis if they are not also manually provided.**
