## Supplementary methods, Figures S1-S4 and Tables S3-S6 for "Developing Del2Phen: a novel phenotype description tool for chromosome deletions"

This manuscript contains the following supplementary information:

**Supplementary Information S1.** Supplementary Methods – page 2
**Figure S1**. Determining the HI-gene overlap between deletions – page 4
**Figure S2.** Inclusion flowchart of cases for the current study – page 5
**Figure S3.** Distribution of deletions along chromosome 6 – page 6
**Figure S4.** Number of individuals with more than *n* connections for a minimum HI-gene similarity threshold – page 7
**Table S1.** Phenotypic features and other clinical characteristics covered by the Chromosome 6 Questionnaire – separate Excel file
**Table S2.** Genes on chromosome 6 that fulfil our definition of HI and are considered HI-genes by Del2Phen – separate Excel file
**Table S3.** Del2Phen's current dominating-effect genes – page 8
**Table S4.** Results of Mann-Whitney U test for the difference in PPV in patients with and without a dominating-effect gene in their deletion for various HI-gene similarity thresholds – page 9
**Table S5.** *p*-values derived from Mann-Whitney U test for differences in PPV in groups that exceed different sizes – page 9
**Table S6.** Example of a clinical description generated by Del2Phen for a patient - without a dominating-effect gene in the deletion – page 10
**README file** for Del2Phen – separate PDF

**Supplementary Information S1.** Supplementary Methods

1. **Data collection within the Chromosome 6 Project and exclusion criteria for the current study**

For this study, Del2Phen uses the data collected through the Chromosome 6 Project and stored in the Chromosome 6 database, which was constructed using the Molgenis toolkit [1]. The project collects genotype and phenotype information from new patients and literature cases with a structural chromosome 6 aberration diagnosed via a microarray or other high-resolution genetic analysis. Phenotype data is collected through an online multilingual questionnaire, the Chromosome 6 Questionnaire, covering a broad range of topics. An overview of these topics is provided by Engwerda et al. [2]. For details on patient recruitment, see our previous publications [3–6].

For the current study, we excluded individuals in the Chromosome 6 database with intragenic deletions, complex rearrangements, or additional genetic aberrations with a possible clinical effect. Small deletions (<0.4 Mb) containing no haploinsufficiency (HI) genes based on current knowledge (see Materials and Methods 2.1. for definition of these genes) were also excluded if: (i) they were described in the Database of Genomic Variants (DGV) (http://dgv.tcag.ca/dgv/app/home) [7] *or* (ii) they were inherited from a healthy parent. Lastly, cases were excluded if an HI-gene was located between the last present and first absent array probe (or vice versa), making it unclear whether that gene was deleted.

2. **Selection of dominating-effect genes**

Dominating-effect genes were chosen based on current knowledge regarding their phenotypic effect as described in the Online Mendelian Inheritance in Men (OMIM) [8,9] and our previous studies [3,4,6]. Only genes with established gene-disease associations (in the case of deletions) or loss-of-function variants known to have a prominent role in the resulting phenotype were selected. GeneHancer was consulted for any known regulatory elements of the chosen dominating-effect genes that lie outside of the gene's chromosome location [10]. Only GeneHancer 'double elite' regions were considered, which are regions that are highly likely to act as regulatory elements of a specific gene. For these regions, Decipher [11], DGV [7] and the Chromosome 6 database were searched for deletions that include only the region of the regulatory element without the target gene. No convincing evidence was found that deletions including any of GeneHancer's double elite regulatory elements for these five genes without the target gene itself have the same clinical effect. Hence, these regions were not incorporated into the tool's parameters.

3. **Del2Phen: additional input and usage**

Del2Phen requires genotype and phenotype information for patients in simple columnar format as input and a reference gene set as a GTF file. The Ensembl gene set is used as reference for gene nomenclature [12]. Gene HI scores (HI, pLI and pHaplo) are provided as additional datasets within the tool [13–15]. Python package MyGene v.3.2.2 is used to homogenise gene IDs across datasets [16–18]. Additional optional inputs include patient and phenotype filter lists, as well as various genotypic similarity parameter settings. In the current study, the patient datasets are not provided externally but are retrieved directly from the Chromosome 6 database.

Del2Phen can be used in one of three ways: (i) as a Python library for interactive or programmatic command-line data exploration, (ii) as a command-line tool to directly output data such as phenotype predictions, or (iii) as an interactive dashboard for live data exploration, parameter alteration and phenotype description. The supplementary README file provides more details on the various ways Del2Phen can be used.

### 4. Phenotype list for determining positive predictive value

Positive predictive value (PPV) was used as an outcome measure for determining the precision of the phenotype descriptions produced by Del2Phen in order to choose its default genotypic similarity settings (see Materials and Methods 2.3.). PPV was calculated using a list of 115 main features (indicated in bold in Table S1) out of the total list of features covered by the Chromosome 6 Questionnaire, in order to prevent double counting of the same feature if both the main and sub-features were known (for example heart defect and ventricular septal defect). Additional exclusion criteria for this list were: (i) main features that were closely related to another feature in the list were omitted, such as “impaired social interactions” and “behavioural diagnosis”, (ii) main features that were highly prevalent in all groups irrespective of chromosome region (such as developmental delay) were also omitted, and (iii) in case of mutually exclusive sub-features (such as “microcephaly” and “macrocephaly”), the sub-features were included instead of the main feature.

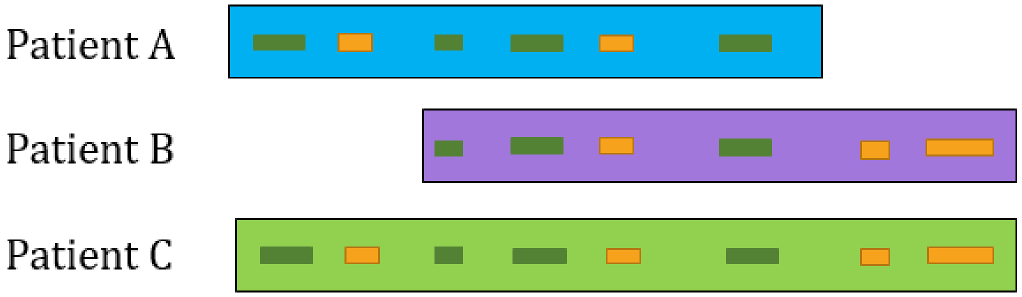


$$J(\mathrm{A, B}) = \frac{A \cap B}{A \cup B} = \frac{1}{1+1+2} = 0.25$$

HI-gene similarity (A, B) = 0.25 x 100 = 25%

$$J(\mathrm{B, C}) = \frac{B \cap C}{B \cup C} = \frac{3}{1+3} = 0.75$$

HI-gene similarity (B, C) = 0.75 x 100 = 75%

**Fig. S1:** Determining the HI-gene overlap between deletions (HI-gene similarity). Orange bars depict the deleted HI-genes. Green bars indicate deleted genes that are not likely to exhibit HI. For the definition of HI-genes, see Materials and Methods 2.1. To determine the HI-gene overlap between deletion pairs, the intersection and union of the sets of deleted HI-genes are calculated, based on which the similarity (Jaccard index) is determined. In the illustrated examples, patients A and B exhibit a 25% HI-gene overlap, while patients B and C share 75% of their HI-gene content. Even though patients A and B have deletions of similar size, the functional genotypic similarity is considerably higher between patients B and C.

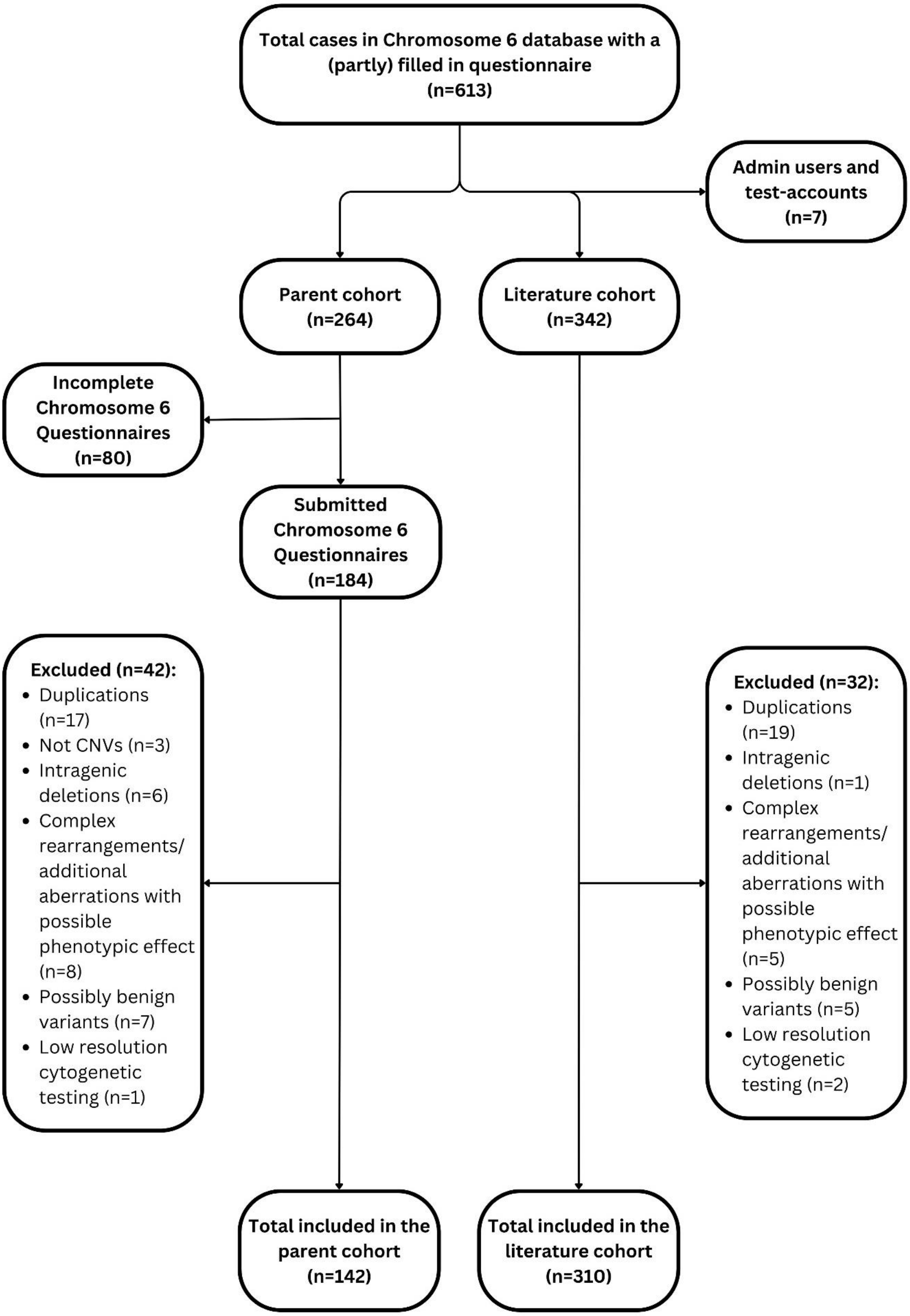


**Fig. S2:** Inclusion flowchart of cases for the current study.

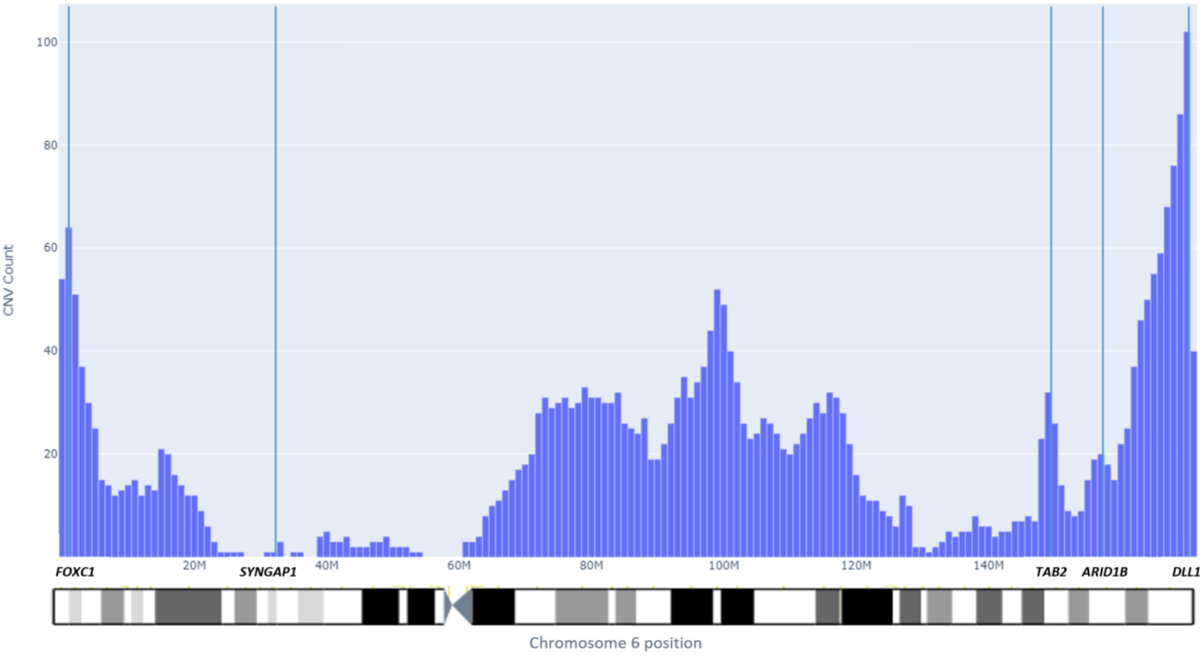


**Fig. S3:** Distribution of deletions along chromosome 6. The figure depicts the frequency (y-axis) at which each base pair (x-axis) on chromosome 6 is deleted in our current cohort. The chromosome is separated into 1 Mb bins. If a bin is at least partially covered by a deletion, it is included in the CNV count. Thin vertical blue lines indicate the location of the five dominating-effect genes (Table S3). This figure shows the unequal distribution of deletions along the chromosome, with a pronounced concentration of deletions towards the terminal 6q end.

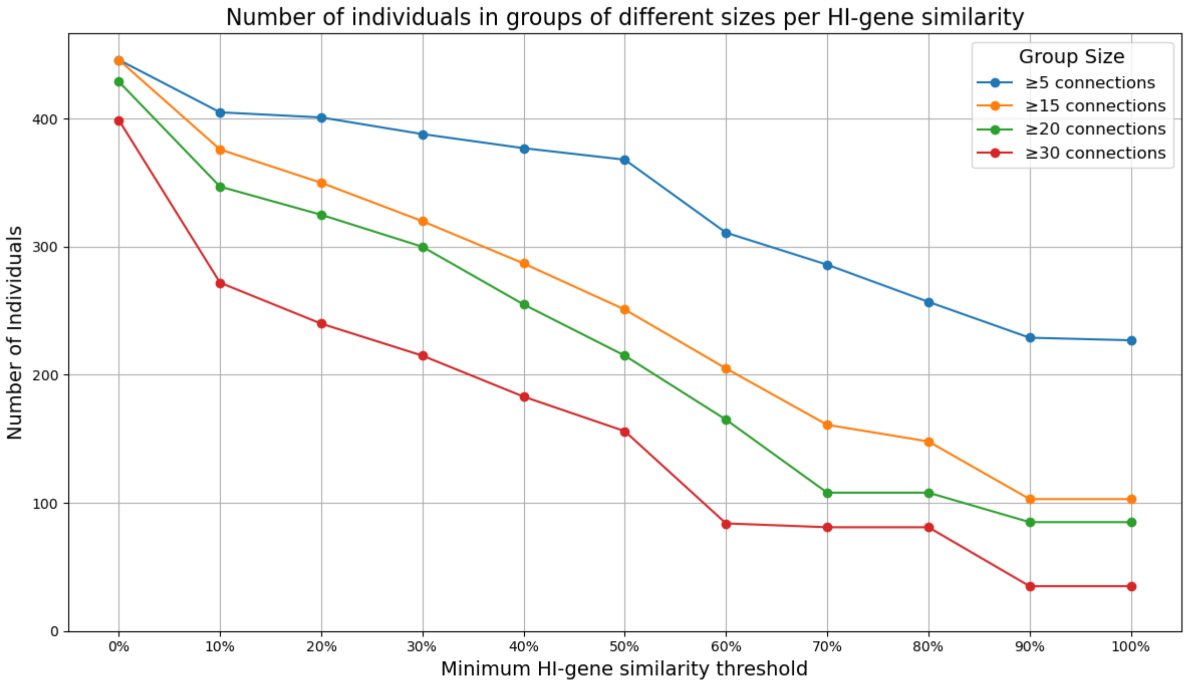


**Fig. S4:** Number of individuals (y-axis) with more than *n* connections for a minimum HI-gene similarity threshold (x-axis). As depicted in this graph, the number of individuals that exceed a certain number of connections (5, 15, 20 or 30) decreases as minimum HI-gene similarity increases.

**Table S3.** Del2Phen's current dominating-effect genes

| Gene symbol and HGNC identifier | Location (in hg19) | HI score | pLI score | pHaplo score | Associated phenotypes[a] |
|---|---|---|---|---|---|
| ***FOXC1*** (HGNC: 3800) | chr6:1610681-1614127 | 9% | 0.95 | 1.00 | Axenfeld-Rieger syndrome (MIM#602482), characterised by ocular anterior segment dysgenesis, hearing loss, distinct craniofacial dysmorphisms, *congenital heart defects and cerebellar abnormalities* [4] |
| ***SYNGAP1*** (HGNC: 11497) | chr6:33387847-33421466 | 19.65% | 1.00 | 0.97 | Developmental delay/intellectual disability, (epileptic) seizures, hypotonia, *behavioural problems, microcephaly and musculoskeletal problems* [19] |
| ***TAB2*** (HGNC: 17075) | chr6:149539777-149732749 | 5.38% | 1.00 | 0.99 | Congenital heart defects mostly of the mitral valve and/or (dilated) cardiomyopathy, short stature, connective tissue abnormalities, distinctive facial features and *mild intellectual disability* [6] |
| ***ARID1B*** (HGNC: 18040) | chr6:157099063-157531913 | 14.17% | 1.00 | 1.00 | Coffin-Siris syndrome 1 (MIM#135900), characterised by developmental or cognitive delay; distinctive hair, nail and skeletal abnormalities; dysmorphic facial features; hypotonia; feeding problems; *poor growth; ophthalmological problems; corpus callosum abnormalities; seizures and behavioural problems* [20,21] |
| ***DLL1*** (HGNC: 2908) | chr6:170591294-170599561 | 4.66% | 1.00 | 1.00 | Developmental delay, behavioural problems, structural brain defects, (spinal) skeletal abnormalities, *hypotonia, seizures, ataxia, eye movement abnormalities and (variable) craniofacial dysmorphisms* [5,22] |

These genes are considered dominating-effect genes based on their predicted HI effect and their highly penetrant and distinctive effect on the phenotype. See Supplementary Information S1 for details on the selection of these genes.

[a]Italicised features are present to a lesser extent.

HI = haploinsufficiency, pHaplo = predicted probability of haploinsufficiency, pLI = probability of loss-of-function intolerance.

**Table S4** Results of Mann-Whitney U test for the difference in PPV in patients with and without a dominating-effect gene in their deletion for various HI-gene similarity thresholds

| Minimum HI-gene similarity used for grouping | Mann-Whitney statistic | p-value* |
|---|---|---|
| **0%** | 29772.5 | **<0.001** |
| **10%** | 23732.0 | **0.006** |
| **20%** | 22429.0 | **0.043** |
| **30%** | 21548.5 | **0.012** |
| **40%** | 19981.0 | **0.033** |
| **50%** | 19200.5 | **0.022** |
| **60%** | 12690.5 | 0.223 |
| **70%** | 10277.0 | 0.275 |
| **80%** | 7862.0 | 0.185 |
| **90%** | 5943.5 | 0.175 |
| **100%** | 5644.5 | 0.28 |

HI = Haploinsufficiency; PPV = positive predictive value. See Materials and Methods 2.1. for definition of HI-genes and section 2.3.1. for definition and calculation of PPV.
* *p*-values in bold indicate a significant difference in PPV between the two groups for the given HI-gene similarity threshold based on α = 0.05.

**Table S5** *p*-values derived from the Mann-Whitney U test for differences in PPV in groups that exceed different sizes

| Minimum HI-gene similarity threshold | *p*-value for PPV in groups < vs. ≥15* | *p*-value for PPV in groups < vs. ≥20* | *p*-value for PPV in groups < vs. ≥30* |
|---|---|---|---|
| 50% | **0.001** | **0.001** | **0.019** |
| 60% | **0.002** | **0.012** | 0.519 |
| 70% | **0.02** | 0.12 | 0.521 |
| 80% | 0.165 | 0.086 | 0.491 |
| 90% | **0.038** | **0.019** | 0.472 |
| 100% | 0.057 | **0.028** | 0.516 |

HI = Haploinsufficiency; PPV = positive predictive value. See Materials and Methods 2.1. for definition of HI-genes and section 2.3.1. for definition and calculation of PPV.
* *p*-values in bold indicate a significant difference in PPV for the given HI-gene similarity threshold based on α = 0.05.

**Table S6** Example of a clinical description generated by Del2Phen for a patient without a dominating-effect gene in the deletion

| Clinical description for patient Id019 | Features present in patient Id019 |
|---|---|
| | *Birth through Caesarean section* |
| Hospitalisation after birth | |
| **Abnormal oral cavity morphology** | |
| **Abnormality of the outer ear** | **Abnormality of the outer ear** |
| Aplasia/Hypoplasia of the external ear | Aplasia/Hypoplasia of the external ear |
| **Abdominal wall defect** | **Abdominal wall defect** |
| Umbilical hernia | Umbilical hernia |
| **Sacral dimple** | |
| **Joint hypermobility** | |
| Generalised joint laxity | |
| **Abnormality of the hand** | **Abnormality of the hand** |
| **Abnormality of the foot** | **Abnormality of the foot** |
| Positional foot deformity | |
| Toe syndactyly | |
| **Feeding difficulties** | |
| Impaired mastication | |
| **Abdominal abnormality** | **Abdominal abnormality** |
| Constipation | Constipation |
| **Abnormality of the kidney** | |
| **Abnormality of the genitalia** | **Abnormality of the genitalia** |
| Cryptorchidism | |
| | Displacement of the urethral meatus |
| **Recurrent infections** | |
| | **Growth hormone deficiency** |
| **Vision problems** | |
| | **Abnormal middle/inner ear morphology** |
| | *Tympanostomy tubes placed* |
| Normal hearing at newborn screening | Normal hearing at newborn screening |
| | ***Hearing impairment*** |
| | Moderate hearing impairment |
| **Brain imaging abnormality** | |
| **Muscular hypotonia** | **Muscular hypotonia** |
| Neurodevelopmental delay | Neurodevelopmental delay |
| Global developmental delay | |
| Delayed gross motor development | Delayed gross motor development |
| Delayed fine motor development | |
| Delayed speech and language development | Delayed speech and language development |
| Receptive language delay | |
| Expressive language delay | Expressive language delay |
| Uses communication aids | |
| **Shy behaviour** | |
| **Social behaviour** | **Social behaviour** |
| **Helpful behaviour** | **Helpful behaviour** |
| Easily upset behaviour | |
| Quiet behaviour | |
| **Phenotype description assessment** | |
| **Total number of features in description (only using the selected feature in bold)** | 38 (18) |
| **Number of features in description seen in patient (only using the selected feature in bold)** | 17 (9) |
| **PPV based on full description (only using the selected feature in bold)** | 0.45 (0.5) |

Clinical description based on a group of 16 individuals sharing a minimum of 70% of their HI-gene content with the index patient. The clinical description in this table is based on all features covered by the Chromosome 6 Questionnaire deriving from closed-ended questions that are present in at least 20% of the connected individuals. Features in bold were used to calculate PPV in this study. Features in italics are those present in the index that were also seen in at least one connected individual but did not meet the prevalence threshold for inclusion in the clinical description.
